# Rare genetic variants in dominant developmental disorder loci cause milder related phenotypes in the general population

**DOI:** 10.1101/2021.12.15.21267855

**Authors:** Rebecca Kingdom, Marcus Tuke, Andrew Wood, Robin N. Beaumont, Timothy Frayling, Michael N. Weedon, Caroline F. Wright

## Abstract

Many rare diseases are known to be caused by deleterious variants in Mendelian genes, however the same variants can also be found in people without the associated clinical phenotypes. The penetrance of these monogenic variants is generally unknown in the wider population, as they are typically identified in small clinical cohorts of affected individuals and families with highly penetrant variants. Here, we investigated the phenotypic effect of rare, potentially deleterious variants in genes and loci that are known to cause monogenic developmental disorders (DD) in a large population cohort. We used UK Biobank to investigate phenotypes associated with rare protein-truncating and missense variants in 599 dominant DD genes using whole exome sequencing data from ∼200,000 individuals, and rare copy number variants overlapping known DD loci using SNP-array data from ∼500,000 individuals. We found that individuals with these likely deleterious variants had a mild DD-related phenotype, including lower fluid intelligence, slower reaction times, lower numeric memory scores and longer pairs matching times compared to the rest of the UK Biobank cohort. They were also shorter, with a higher BMI and had significant socioeconomic disadvantages, being less likely to be employed or be able to work, and having a lower income and higher deprivation index. Our findings suggest that many monogenic DD genes routinely tested within paediatric genetics have intermediate penetrance and may cause lifelong milder, sub-clinical phenotypes in the general adult population.

## INTRODUCTION

Many rare diseases are caused by deleterious variants in thousands of monogenic disease genes^1^. However, not all individuals with these variants share the same clinical phenotypes; some don’t appear to be affected at all, whereas others are very severely affected^2^. Monogenic variants can display different effects in different individuals^3^. The range of phenotypes caused by deleterious variants in the same gene can be explained by pleiotropy, incomplete penetrance and variable expressivity^4^. Penetrance (i.e. whether an individual with a disease-causing genotype displays the corresponding clinical phenotype) is generally binary; either a variant is penetrant and causes the clinical phenotype associated with that genotype, or it is not^2,5^. In contrast, variable expressivity (i.e. the range of phenotypes that can be observed in affected individuals) is generally continuous, e.g. from mild to severe^6^. As most disease-causing monogenic variants have been identified through small clinical cohorts, including families with multiple affected individuals, penetrance of these variants is often over-estimated. Investigating the effect of these variants in the general population is therefore important to give a more accurate view of the penetrance in clinically unselected individuals and families. It has been suggested that many of the primary symptoms of rare disease are actually extremes of normally distributed phenotypes in the general population^1,7^. Large, well genotyped population cohorts give us the ability to investigate the spectrum of phenotypes of individuals with variants in known monogenic disease-causing genes. Phenotypic heterogeneity and variability are a major concern for rare Mendelian disorders, where they can lead to incorrect or delayed diagnoses^8,9^.

Many severe developmental disorders (DD) manifest from birth or early childhood and are caused by rare damaging variants in around 2,000 genes and loci^10^. Pathogenic variants in these genes have been identified primarily through phenotype-led clinical studies of affected individuals and families^4^. Due to extensive genetic and phenotypic heterogeneity, large multigene panels are increasingly being used for diagnostic testing, often through panel-based virtual analysis of whole exome or genome sequence data. However, little is known about what effect, if any, deleterious variants in these genes have on adults in the general population or their life-long implications. In this study, using genetic and phenotypic data from UK Biobank (UKB)^11^, we investigated whether adults with rare deleterious variants in genes and loci known to cause autosomal dominant forms of DD have any developmentally-relevant phenotypes.

## MATERIALS AND METHODS

### UK Biobank cohort

UKB is a population-based cohort from the UK with deep phenotyping data and genetic data for around 500,000 individuals aged 40-70 years at recruitment. Individuals provided a variety of information via self-report questionnaires, cognitive and anthropometric measurements, and Hospital Episode Statistics (HES) including ICD9 and ICD10 codes. Genotypes for single nucleotide polymorphisms (SNPs) were generated using the Affymetrix Axiom UK Biobank array (∼450,000 individuals) and the UK BiLEVE array (∼50,000 individuals). This dataset underwent extensive central quality control (http://biobank.ctsu.ox.ac.uk). A subset of ∼200,000 individuals also underwent whole exome sequencing (WES) using the IDT xGen Exome Research Panel v1.0; this dataset was made available for research in October 2020. Detailed sequencing and variant detection methodology for UKB is available at https://biobank.ctsu.ox.ac.uk/showcase/label.cgi?id=170. The UKB resource was approved by the UK Biobank Research Ethics Committee and all participants provided written informed consent to participate.

### Gene selection

We used the clinically curated Developmental Disorders Genotype-to-Phenotype Database (DDG2P) to select genes known to cause monogenic DD. The database (accessed on 27 November 2020) was constructed from published literature and provides information relating to genes, variants and phenotypes associated with DDs, including mode of inheritance and mechanism of pathogenicity^10^. We initially included all genes that had been annotated as a “confirmed” or “probable” causes of autosomal dominant DD (n=599). Further subsets of these genes were selected for sensitivity analyses, including: a panel of 384 genes that are known to cause DD through a loss-of-function (LoF) mechanism; a more stringent panel of 125 of these haploinsufficiency genes that were significantly enriched for damaging *de novo* loss-of-function (LoF) mutations in a recent analysis of 31,058 DD probands^12^; and a small panel of 25 clinically well-established genes with >30 likely pathogenic *de novo* LoF mutations in the same study^12^ (see **Figure 1** and **Supplementary Table 1**).

**Figure 1.**
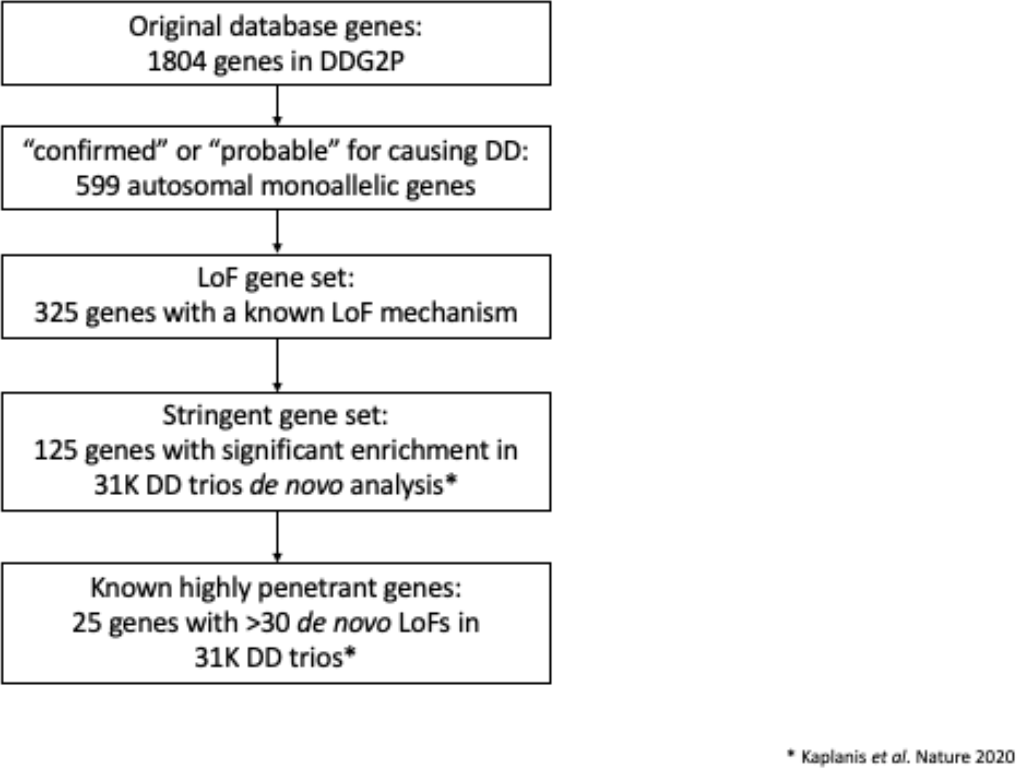
Flow diagram outlining selection process for DD genes in each subset that were used for analysis. DDG2P = Developmental Disorders Genotype-to-Phenotype database; DD = Developmental Disorder; LoF = Loss of Function variants; 31K DD trios = 31,058 parent-offspring families with developmental disorders (Kaplanis et al. 2020).

### Variant selection

To investigate the penetrance of likely deleterious single nucleotide variants (SNVs) and insertions/deletions (indels) in known autosomal dominant DD genes, we used WES data from 200,632 individuals in UKB to identify individuals with a rare SNVs and/or indels in any of these genes. For most of our analyses, rare was defined as any variant that occurred in 5 or fewer individuals in the UKB cohort; we also investigated the effect of changing this threshold to n=1, n=10, n=50 and n=100 individuals. We included variants that had individual and variant missingness <10%, minimum read depth of 7 for SNVs and 10 for indels, and at least one sample per site passed the allele balance threshold > 15% for SNVs and 20% for indels. We selected three functional classes of variant in canonical transcripts based on annotation by the Ensembl Variant Effect Predictor^13^:

1. *Likely deleterious LoF variants:* we defined a LoF variant as one that is predicted to cause a premature stop, a frameshift, or abolish a canonical splice site; only those variants deemed to be high confidence by the Loss-Of-Function Transcript Effect Estimator (LOFTEE) were retained (https://github.com/konradjk/loftee).
2. *Likely deleterious missense variants:* missense variants with a Combined Annotation Dependent Depletion (CADD)^14^ score ≥ 30.
3. *Likely benign synonymous variants*.

Individuals with variants in group (1) were excluded from groups (2) and (3); individuals with variants in group (2) were excluded from group (3). Following variant selection, one gene (*DNMT3A*) was removed from further analysis as the variants in this gene – which is known to be strongly linked with blood cancer^15^ – had a significantly lower allele balance, suggesting substantial somatic mosaicism (see **Supplementary Figure 1**). LoF variants in the most stringent 25 gene subset were visually confirmed using the Integrative Genomics Viewer (IGV).

To investigate the penetrance of multigenic copy number variants (CNVs) overlapping known DD loci, we used SNP-array data from 488,377 genotyped individuals in UKB and PennCNV^16^ (version 1.0.4) to detect multigenic CNVs overlapping 69 published CNVs strongly associated with developmental delay^17,18^. Log R ratio (LRR) and B-allele frequency (BAF) values for 805,426 genome-wide SNP probe sets were provided by UKB, and an in-house script was used to convert these data to PennCNV input signal files. The PennCNV Hidden Markov Model (HMM) transition matrix was trained using 250 random UK Biobank samples using PennCNV-train. Population Frequency B Allele reference data (PFB) were generated using 1,000 random UK Biobank samples. PennCNV-test was then used to detect regions in a duplication or deletion state in LRR/BAF Hidden Markov Model (HMM) with the generated PFB and transition matrix. An individual was classified as having a multigenic DD deletion or duplication if the region detected using PennCNV reciprocally intersected the published region by at least 50%. We plotted LRR/BAF data for each call in each of these regions, and carried out visual inspection of each event, and false positives and single gene CNVs were excluded. A list of included CNVs included is provided in **Supplementary Table 2**.

### Statistical analysis

We performed both individual gene and gene panel burden tests across our different gene subsets. We grouped individuals into one of five groups depending upon the type of variant they carried (LoF, missense or synonymous variant in one or more autosomal dominant DD genes; or deletion or duplication overlapping published DD multigenic CNVs). Association tests were limited to individuals in UKB with genetically defined European ancestry that were unrelated up to third-degree relationship (184,142 with WES data; 380,029 with SNP-array data) and were controlled for age, sex, recruitment centre and 40 principal components. Variant burden association tests in gene panels and multigenic CNVs were performed using STATA (version 16.0), using linear regression for continuous phenotypes and logistic regression for the binary phenotypes. Associations were tested between each group of individuals and other individuals in the UKB cohort without any of the classes of rare variation defined above. Information from HES codes, self-report questionnaires and cognitive tests taken at recruitment was used for the phenotypic information. Associations were tested for 22 UKB phenotypes selected based on their likely relevance to developmental disorders, including:

- *Medical*: epilepsy (self-reported or ICD10 codes G40); ever reported a mental health issue (self-reported through questionnaire); diagnosed with “Child DD” (including mental retardation (ICD10 codes F70-73), epilepsy (G40), developmental disorders (F80-84) and congenital malformations (Q0-99)); or diagnosed “Adult DD” (including schizophrenia, (self-reported or ICD10 codes F20-29) and bipolar disorder (self-reported or ICD10 codes F30-F39)).
- *Reproductive*: infertility, number of pregnancies, number of stillbirths, number of children fathered.
- *Physical*: height, body mass index (BMI) (inverse normalised).
- *Cognitive*: fluid intelligence (Field ID: 20016), reaction time (inverse normalised, Field ID: 20023), pairs matching score (Field ID: 20131), numeric memory (inverse normalised, Field ID: 20240), age left education, number of years in education, has a degree.
- *Socioeconomic*: in employment, unable to work (both Field ID: 6142), income (Field ID: 738), Townsend Deprivation Index (TDI) (Field ID: 189).

## RESULTS

### Many individuals in UKB carry rare, deleterious variants in autosomal dominant DD genes

Although each gene individually accounts for extremely rare forms of DD and has a small burden of rare deleterious variants, together they account for a large portion of DD diagnoses and have a surprisingly high burden of rare deleterious variants in UKB. In 184,477 unrelated European individuals with WES data in UKB and across 599 autosomal dominant DD genes: 9103 individuals carry a rare (n≤5) LoF variant, 9846 individuals carry a rare missense variant with CADD>30, and 79,959 individuals carry a rare synonymous variant. As the gene panel becomes smaller and more stringent, the burden of rare deleterious variants decreases; for example, 3602, 1327 and 167 individuals in UKB carry rare LoF variants in smaller more stringent subsets of 384, 125 and 25 monogenic DD genes, respectively (**Figure 1**). In 450,274 individuals with SNP-array data in UKB and across 69 known DD loci, 4922 individuals carry large deletions and 7054 individuals carry large duplications.

### Individuals in UKB with rare, deleterious variants in known DD loci display milder DD-related phenotypes

We performed gene panel (including 599 autosomal dominant genes) and multigenic copy number (including 53 deletions/duplications syndromes) burden tests for 22 traits in UKB selected to be of relevance (in adults) to developmental phenotypes. Bonferroni-corrected significant associations were found across most phenotypes in individuals carrying likely damaging variants compared with the rest of the UKB cohort (**Table 1** and **Figure 2**). Individuals carrying these variants generally had lower cognitive performance than the rest of the cohort, with reduced fluid intelligence (LoF group beta: −1.059), slower reaction times (LoF group beta: +0.043), lower numeric memory scores (LoF group beta: −0.068) and longer pairs matching times (LoF group beta: +0.122). They also completed fewer years in education, left education at an earlier age and were less likely to have a degree. Medically, individuals were more likely to have reported a mental health issue or been diagnosed with either a childhood DD (including mild-severe mental retardation, epilepsy, autism, ADHD, and congenital malformations) or adult DD (including schizophrenia and bipolar disorder). Individuals were also more likely to be shorter, have a higher BMI and have had fewer children (though the latter association was only significant in men). Individuals also had significant socioeconomic disadvantages, being less likely to be employed or be able to work, having a lower income and a higher deprivation index (TDI). Across all phenotypes tested, we observed a trend corresponding to the likely deleteriousness of the variants; the largest effect was generally observed in the group of individuals with multigenic deletions, followed by multigenic duplications, then LoF variants and finally missense variants in one (or more) DD genes. In contrast, individuals with only rare synonymous variants in DD genes showed no statistically significant difference in any phenotype compared to the remainder of the cohort, as expected for likely benign variants.

**Table 1.**
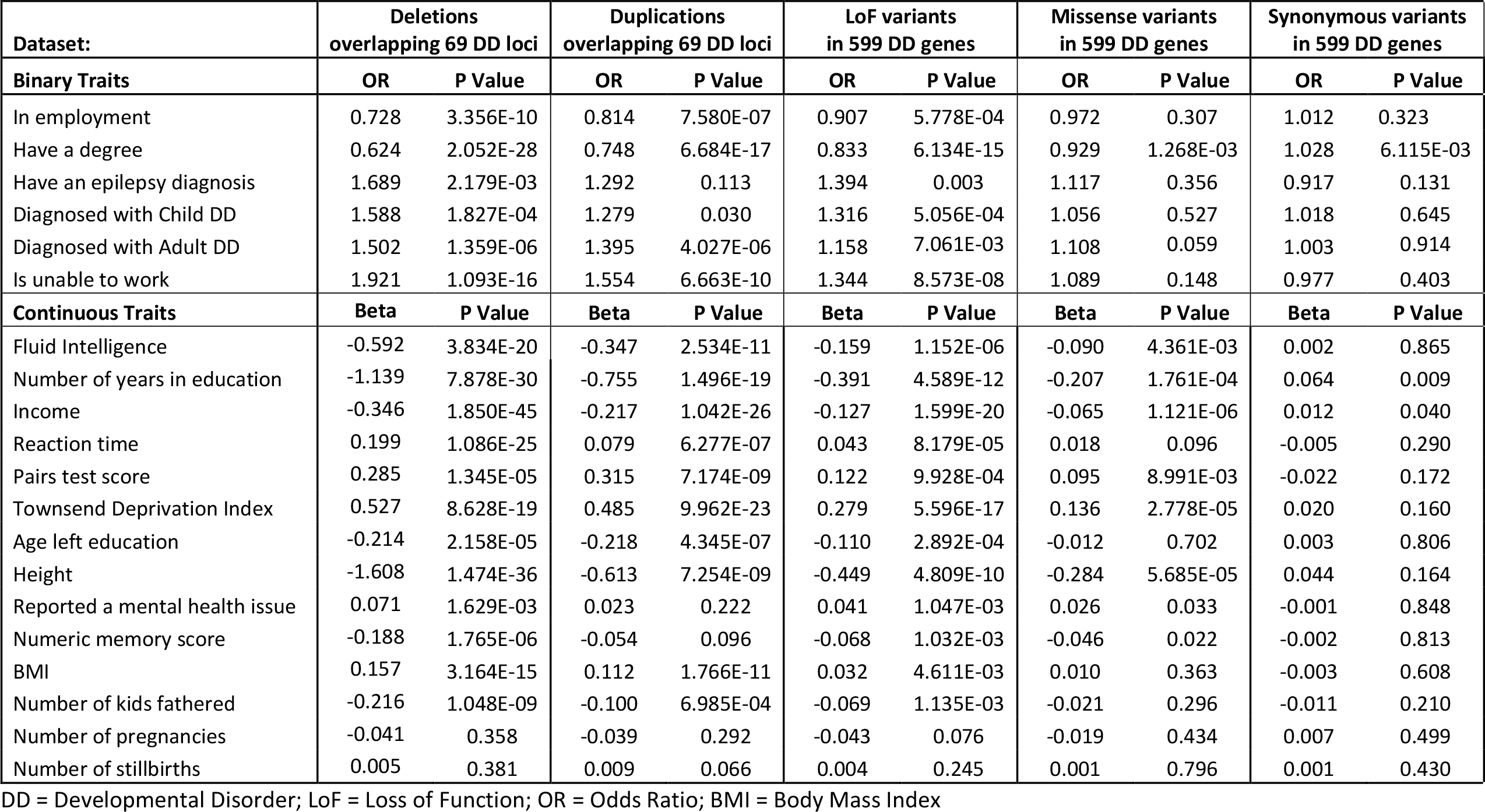
Gene panel association tests for 22 phenotypes tested in individuals in UK Biobank carrying deletions or duplications overlapping 69 known DD syndromic loci, or rare (n≤5) LoF, missense (CADD>30) or synonymous variants in any of 599 known autosomal dominant DD genes.

| Dataset: | Deletions<br>overlapping 69 DD loci |  | Duplications<br>overlapping 69 DD loci |  | LoF variants<br>in 599 DD genes |  | Missense variants<br>in 599 DD genes |  | Synonymous variants<br>in 599 DD genes |  |
| --- | --- | --- | --- | --- | --- | --- | --- | --- | --- | --- |
| Binary Traits | OR | P Value | OR | P Value | OR | P Value | OR | P Value | OR | P Value |
| In employment | 0.728 | 3.356E-10 | 0.814 | 7.580E-07 | 0.907 | 5.778E-04 | 0.972 | 0.307 | 1.012 | 0.323 |
| Have a degree | 0.624 | 2.052E-28 | 0.748 | 6.684E-17 | 0.833 | 6.134E-15 | 0.929 | 1.268E-03 | 1.028 | 6.115E-03 |
| Have an epilepsy diagnosis | 1.689 | 2.179E-03 | 1.292 | 0.113 | 1.394 | 0.003 | 1.117 | 0.356 | 0.917 | 0.131 |
| Diagnosed with Child DD | 1.588 | 1.827E-04 | 1.279 | 0.030 | 1.316 | 5.056E-04 | 1.056 | 0.527 | 1.018 | 0.645 |
| Diagnosed with Adult DD | 1.502 | 1.359E-06 | 1.395 | 4.027E-06 | 1.158 | 7.061E-03 | 1.108 | 0.059 | 1.003 | 0.914 |
| Is unable to work | 1.921 | 1.093E-16 | 1.554 | 6.663E-10 | 1.344 | 8.573E-08 | 1.089 | 0.148 | 0.977 | 0.403 |
| Continuous Traits | Beta | P Value | Beta | P Value | Beta | P Value | Beta | P Value | Beta | P Value |
| Fluid Intelligence | -0.592 | 3.834E-20 | -0.347 | 2.534E-11 | -0.159 | 1.152E-06 | -0.090 | 4.361E-03 | 0.002 | 0.865 |
| Number of years in education | -1.139 | 7.878E-30 | -0.755 | 1.496E-19 | -0.391 | 4.589E-12 | -0.207 | 1.761E-04 | 0.064 | 0.009 |
| Income | -0.346 | 1.850E-45 | -0.217 | 1.042E-26 | -0.127 | 1.599E-20 | -0.065 | 1.121E-06 | 0.012 | 0.040 |
| Reaction time | 0.199 | 1.086E-25 | 0.079 | 6.277E-07 | 0.043 | 8.179E-05 | 0.018 | 0.096 | -0.005 | 0.290 |
| Pairs test score | 0.285 | 1.345E-05 | 0.315 | 7.174E-09 | 0.122 | 9.928E-04 | 0.095 | 8.991E-03 | -0.022 | 0.172 |
| Townsend Deprivation Index | 0.527 | 8.628E-19 | 0.485 | 9.962E-23 | 0.279 | 5.596E-17 | 0.136 | 2.778E-05 | 0.020 | 0.160 |
| Age left education | -0.214 | 2.158E-05 | -0.218 | 4.345E-07 | -0.110 | 2.892E-04 | -0.012 | 0.702 | 0.003 | 0.806 |
| Height | -1.608 | 1.474E-36 | -0.613 | 7.254E-09 | -0.449 | 4.809E-10 | -0.284 | 5.685E-05 | 0.044 | 0.164 |
| Reported a mental health issue | 0.071 | 1.629E-03 | 0.023 | 0.222 | 0.041 | 1.047E-03 | 0.026 | 0.033 | -0.001 | 0.848 |
| Numeric memory score | -0.188 | 1.765E-06 | -0.054 | 0.096 | -0.068 | 1.032E-03 | -0.046 | 0.022 | -0.002 | 0.813 |
| BMI | 0.157 | 3.164E-15 | 0.112 | 1.766E-11 | 0.032 | 4.611E-03 | 0.010 | 0.363 | -0.003 | 0.608 |
| Number of kids fathered | -0.216 | 1.048E-09 | -0.100 | 6.985E-04 | -0.069 | 1.135E-03 | -0.021 | 0.296 | -0.011 | 0.210 |
| Number of pregnancies | -0.041 | 0.358 | -0.039 | 0.292 | -0.043 | 0.076 | -0.019 | 0.434 | 0.007 | 0.499 |
| Number of stillbirths | 0.005 | 0.381 | 0.009 | 0.066 | 0.004 | 0.245 | 0.001 | 0.796 | 0.001 | 0.430 |

DD = Developmental Disorder; LoF = Loss of Function; OR = Odds Ratio; BMI = Body Mass Index

**Figure 2.**
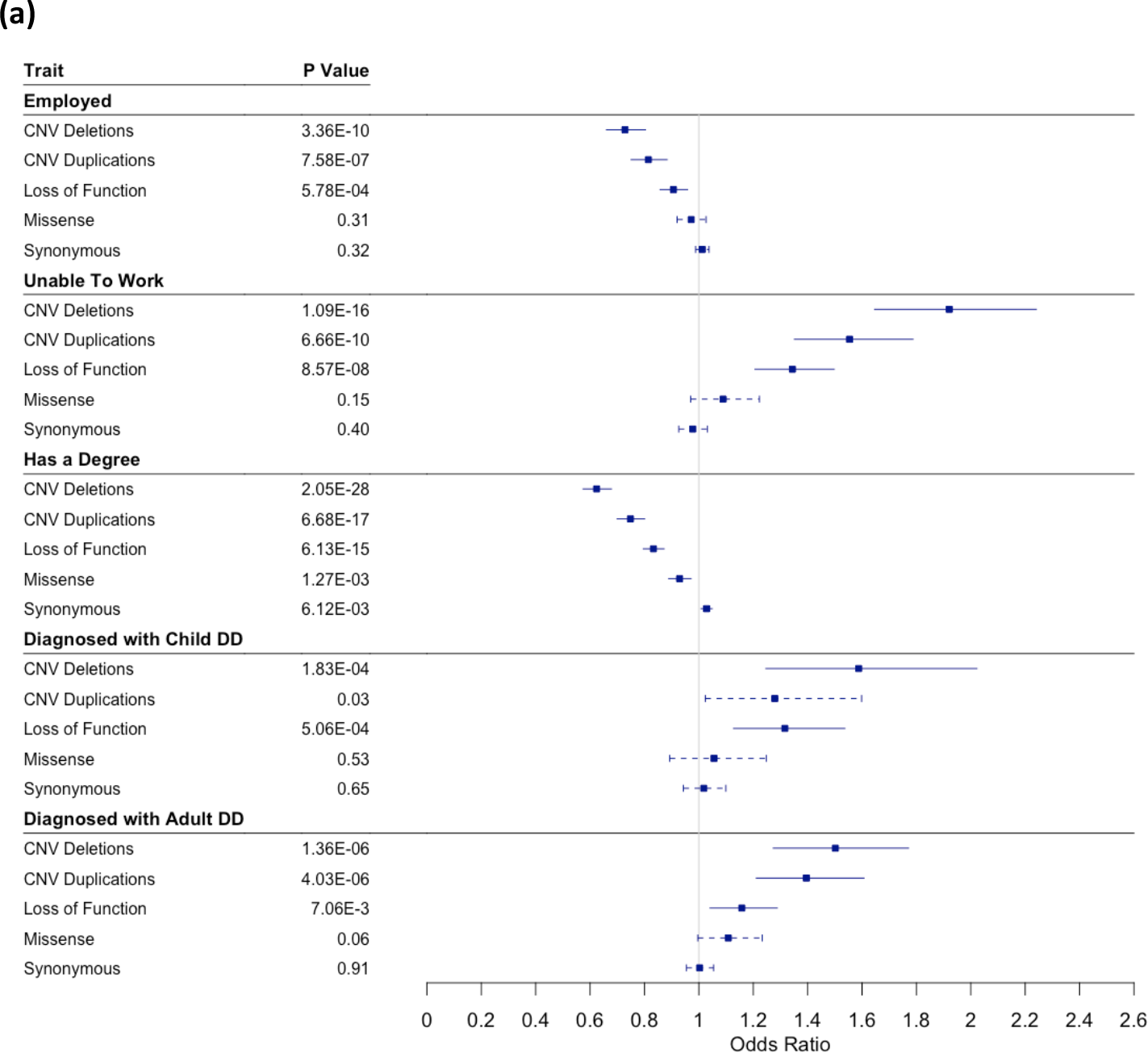
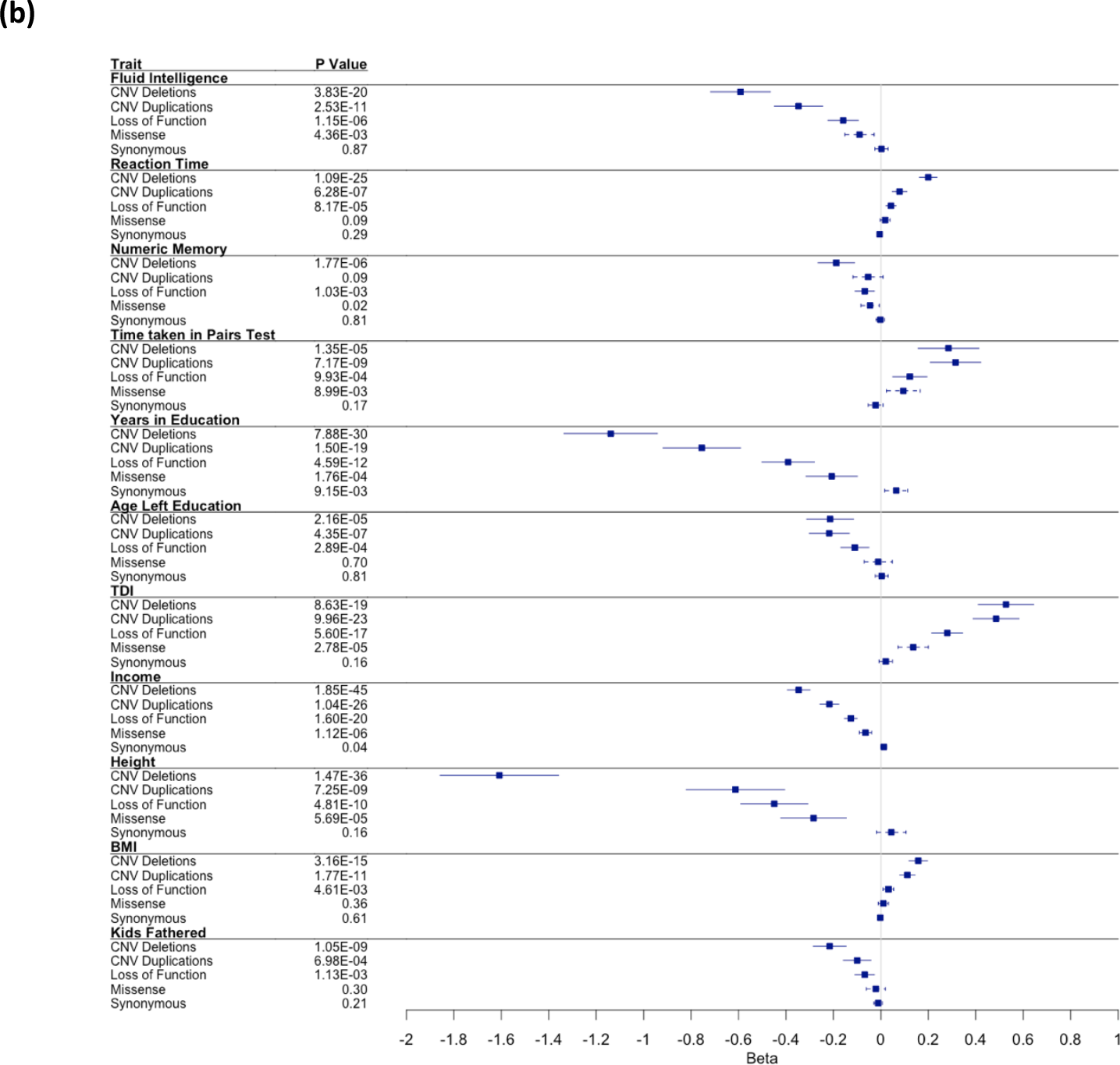
Summary of gene panel association tests for carriers of likely deleterious variants in known autosomal dominant DD loci for **(a)** binary traits **(b)** continuous traits. Associations are shown for individuals carrying deletions or duplications overlapping 69 known DD syndromic loci, or rare (n≤5) LoF, missense (CADD>30) or synonymous variants in any of 599 known autosomal dominant DD genes, compared with the remaining unrelated white Europeans in UKB. Unbroken lines = below Bonferroni-corrected p-value; dashed lines = above Bonferroni-corrected p-value.

### Potentially damaging LoF variants were found even in clinically well-established DD genes previously thought to be highly penetrant

We repeated our association analysis with smaller, more stringent, subsets of 325, 125 and 25 known DD genes (**Figure 1**). Interestingly, even within the most stringent subset of 25 genes that are thought to be highly penetrant causes of DD via haploinsufficiency, with >30 *de novo* LoF mutations identified in 31,058 DD probands^12^, we were able to identify 167 individuals in UKB who had a high confidence LoF variant in one of these genes. We observed similar trends to the full 599 gene panel for LoF variants in smaller subsets of genes cause DD by haploinsufficiency, with the group overall exhibiting mild DD-related phenotypes, though the results were less significant due to the smaller number of individuals carrying likely LoF variants (**Supplementary Table 3**). Nonetheless, a Bonferroni-corrected significant result was seen across all gene subsets for shorter stature, reduced chance of having a degree and increased TDI; lower fluid intelligence, lower income, higher BMI and an increased chance of being diagnosed with a child DD also remained nominally significant even in the 25 gene subset. We also performed single gene burden testing but were underpowered to find any significant associations for most genes due to the small number of individuals and likely mild phenotypic effects in UKB. Interestingly, despite previously reaching genome-wide significance for enrichment of damaging *de novo* mutations, *MIB1* had the largest number of individuals carrying likely LoF variants in UKB (n=260), more than the 25 most stringent genes combined, but showed no associations with any DD-related phenotypes. The gene also has almost double the number of LoF variants observed versus expected in gnomAD^19^, and thus appears to be remarkably unconstrained and thus may not be a true haploinsufficient DD gene.

### Rarer LoF variants have a larger effect than more common LoF variants

We investigated the effect of minor allele count (MAC) on the phenotypic effect of LoF variants in our largest gene panel (599 autosomal dominant DD genes). Specifically we performed association tests with 16 DD-related traits that were significant in the previous analysis for groups of individuals with rare LoF variants in these genes that were present in just a single individual in UKB, compared with variants seen 5, 10, 50 or 100 or fewer times (**Figure 3**). The group of individuals who had the rarest variants (MAC=1) had the largest phenotypic effect change compared to the rest of the cohort, though the results were generally not significant due to low numbers. However, across the phenotypes tested, both the effect size and the p-value decreased as the MAC increased, suggesting either that that more common variants have a milder effect on phenotype, or that more common variants are benign and are simply diluting the effect of rare pathogenic variants. No difference was observed between the effect of LoF variants in the first and second half of genes. In addition, 295 individuals had LoF variants that were previously classified as “likely pathogenic” or “pathogenic” in ClinVar, but no significant difference was detectable in their phenotypes compared with the remainder of the group who also had LoF variants.

**Figure 3.**
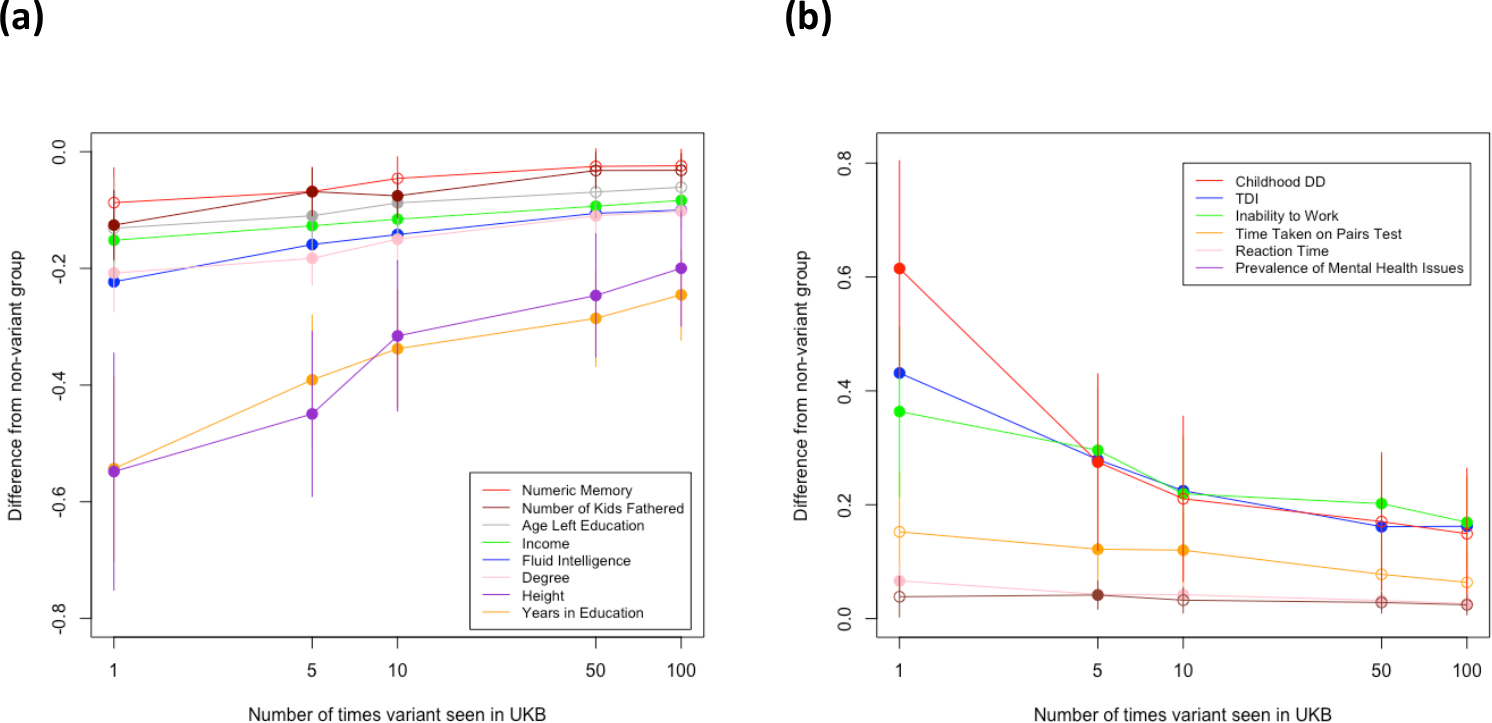
Change in phenotype associations for individuals with a LoF variant in 599 known autosomal dominant DD genes versus different minor allele counts. Associations are grouped by whether the effect of MAC=1 LoF variants either **(a)** decreases or **(b)** increases the phenotype.

## DISCUSSION

We have shown that rare, potentially damaging variants in genes and loci known to cause autosomal dominant DD are present in adults in UK Biobank and result in a mild developmental phenotype. Individuals carrying these variants have notably reduced cognitive abilities and a lower socioeconomic status. Gene panel association tests suggest a strong and consistent trend for increasing phenotypic effects with rarer and more damaging variants. Our findings are consistent with similar studies^4,20–24^ showing reduced penetrance of rare damaging variants in monogenic forms of DD in clinically unselected population cohorts.

We note that the variants identified in UKB are not necessarily the same ones that have been identified previously in clinical cases, and indeed very few of those we identified had previously been annotated in ClinVar^25^. Nonetheless, our findings were robust when limited to likely LoF variants in a subset of 384 DD genes that act via a haploinsufficiency mechanism. The fact that our findings are robust to smaller, more stringent subsets of genes also suggests that the effect cannot simply be explained by a subset of low penetrance (or non-causal) DD genes. Furthermore, rare predicted LoF variants were found in individuals in genes that were thought to be fully or nearly fully penetrant causes of very well-established developmental syndromes, but without the full clinical phenotype that would be expected, suggesting that there is a range of penetrance and expressivity in the general population.

Despite the large size of UKB, we were limited by the number of individuals carrying rare damaging variants in these genes, which meant some of our analyses were under-powered to show a significant effect. We were also limited by the clinical and phenotypic data available on these individuals, all of whom were over 40 years of age at recruitment. Nonetheless, when found in an appropriate clinical paediatric setting, rare damaging variants in these genes are widely considered diagnostic for DD, and thus they might not be expected to be present in a population cohort. Our results suggest that, although the penetrance of variants across these genes is lower than would be expected from previous clinical studies, they do still exert a phenotypic effect on adults in the general population who are nonetheless healthy enough, and have sufficient capacity, to volunteer to participate in a biobank.

Genes and loci that cause monogenic DD have historically been identified almost exclusively through clinical cohorts of affected children and families, and their effect on adults in the general population has not previously been evaluated. While clinical studies may overestimate the penetrance of such rare variants, population cohorts like UKB are likely to underestimate the penetrance, due to ascertainment bias towards healthy individuals^26^. The penetrance and expressivity of variants in these genes could be affected by a number of different modifiers, including genetic variants in other genes, regulatory variants affecting gene expression, somatic mosaicism, and accumulated environmental factors^5^. The latter is particularly relevant when considering the effect of damaging variants in DD genes on adults. It is interesting to note that, unlike most traits, the heritability of intelligence (i.e. general cognitive ability) increases dramatically with age^27^, suggesting a major role for gene-environment interactions as individuals become better able to select, modify and optimise their environment. Further research is needed into the penetrance of rare, damaging variants in the general population using larger datasets, which may allow modifiers to be investigated to help explain why some individuals are more severely affected by particular genetic conditions than others.

## Data Availability

All data used in the present study are available through UK Biobank.

https://www.ukbiobank.ac.uk/

## Acknowledgements

This research has been conducted using the UK Biobank Resource under Application Number 49847. The authors would like to acknowledge funding from the University of Exeter and the MRC (MR/T00200X/1), and the use of the University of Exeter High-Performance Computing (HPC) facility in carrying out this work.

## References

1. Niemi, M.E.K., Martin, H.C., Rice, D.L., Gallone, G., Gordon, S., Kelemen, M., McAloney, K., McRae, J., Radford, E.J., Yu, S., et al. (2018). Common genetic variants contribute to risk of rare severe neurodevelopmental disorders. Nature 562, 268–271.

2. Gruber, C., and Bogunovic, D. (2020). Incomplete penetrance in primary immunodeficiency: a skeleton in the closet. Hum. Genet. 139, 745–757.

3. Oetjens, M.T., Kelly, M.A., Sturm, A.C., Martin, C.L., and Ledbetter, D.H. (2019). Quantifying the polygenic contribution to variable expressivity in eleven rare genetic disorders. Nat. Commun. 10, 4897.

4. Wright, C.F., West, B., Tuke, M., Jones, S.E., Patel, K., Laver, T.W., Beaumont, R.N., Tyrrell, J., Wood, A.R., Frayling, T.M., et al. (2019). Assessing the Pathogenicity, Penetrance, and Expressivity of Putative Disease-Causing Variants in a Population Setting. Am. J. Hum. Genet. 104, 275–286.

5. Cooper, D.N., Krawczak, M., Polychronakos, C., Tyler-Smith, C., and Kehrer-Sawatzki, H. (2013). Where genotype is not predictive of phenotype: towards an understanding of the molecular basis of reduced penetrance in human inherited disease. Hum. Genet. 132, 1077–1130.

6. Shawky, R.M. (2014). Reduced penetrance in human inherited disease. Egypt. J. Med. Hum. Genet. 15, 103–111.

7. Castel, S.E., Cervera, A., Mohammadi, P., Aguet, F., Reverter, F., Wolman, A., Guigo, R., Iossifov, I., Vasileva, A., and Lappalainen, T. (2018). Modified penetrance of coding variants by cis-regulatory variation contributes to disease risk. Nat. Genet. 50, 1327–1334.

8. Rahit, K.M.T.H., and Tarailo-Graovac, M. (2020). Genetic Modifiers and Rare Mendelian Disease. Genes 11, 239.

9. Maya, I., Sukenik-Halevy, R., Basel-Salmon, L., and Sagi-Dain, L. (2020). Ten points to consider when providing genetic counseling for variants of incomplete penetrance and variable expressivity detected in a prenatal setting. Acta Obstet. Gynecol. Scand. 99, 1427–1429.

10. Wright, C.F., Fitzgerald, T.W., Jones, W.D., Clayton, S., McRae, J.F., van Kogelenberg, M., King, D.A., Ambridge, K., Barrett, D.M., Bayzetinova, T., et al. (2015). Genetic diagnosis of developmental disorders in the DDD study: a scalable analysis of genome-wide research data. Lancet Lond. Engl. 385, 1305–1314.

11. Bycroft, C., Freeman, C., Petkova, D., Band, G., Elliott, L.T., Sharp, K., Motyer, A., Vukcevic, D., Delaneau, O., O’Connell, J., et al. (2018). The UK Biobank resource with deep phenotyping and genomic data. Nature 562, 203–209.

12. Kaplanis, J., Samocha, K.E., Wiel, L., Zhang, Z., Arvai, K.J., Eberhardt, R.Y., Gallone, G., Lelieveld, S.H., Martin, H.C., McRae, J.F., et al. (2020). Evidence for 28 genetic disorders discovered by combining healthcare and research data. Nature 586, 757–762.

13. Thormann, A., Halachev, M., McLaren, W., Moore, D.J., Svinti, V., Campbell, A., Kerr, S.M., Tischkowitz, M., Hunt, S.E., Dunlop, M.G., et al. (2019). Flexible and scalable diagnostic filtering of genomic variants using G2P with Ensembl VEP. Nat. Commun. 10, 2373.

14. Rentzsch, P., Witten, D., Cooper, G.M., Shendure, J., and Kircher, M. (2019). CADD: predicting the deleteriousness of variants throughout the human genome. Nucleic Acids Res. 47, D886–D894.

15. Qi, H., Dong, C., Chung, W.K., Wang, K., and Shen, Y. (2016). Deep genetic connection between cancer and developmental disorders. Hum. Mutat. 37, 1042–1050.

16. Wang, K., Li, M., Hadley, D., Liu, R., Glessner, J., Grant, S.F.A., Hakonarson, H., and Bucan, M. (2007). PennCNV: an integrated hidden Markov model designed for high-resolution copy number variation detection in whole-genome SNP genotyping data. Genome Res. 17, 1665–1674.

17. Cooper, G.M., Coe, B.P., Girirajan, S., Rosenfeld, J.A., Vu, T.H., Baker, C., Williams, C., Stalker, H., Hamid, R., Hannig, V., et al. (2011). A copy number variation morbidity map of developmental delay. Nat. Genet. 43, 838–846.

18. Coe, B.P., Witherspoon, K., Rosenfeld, J.A., van Bon, B.W.M., Vulto-van Silfhout, A.T., Bosco, P., Friend, K.L., Baker, C., Buono, S., Vissers, L.E.L.M., et al. (2014). Refining analyses of copy number variation identifies specific genes associated with developmental delay. Nat. Genet. 46, 1063–1071.

19. MIB1 | gnomAD v2.1.1 | gnomAD.

20. Kendall, K.M., Bracher-Smith, M., Fitzpatrick, H., Lynham, A., Rees, E., Escott-Price, V., Owen, M.J., O’Donovan, M.C., Walters, J.T.R., and Kirov, G. (2019). Cognitive performance and functional outcomes of carriers of pathogenic copy number variants: analysis of the UK Biobank. Br. J. Psychiatry 214, 297–304.

21. Crawford, K., Bracher-Smith, M., Owen, D., Kendall, K.M., Rees, E., Pardiñas, A.F., Einon, M., Escott-Price, V., Walters, J.T.R., O’Donovan, M.C., et al. (2019). Medical consequences of pathogenic CNVs in adults: analysis of the UK Biobank. J. Med. Genet. 56, 131–138.

22. Gardner, E.J., Neville, M.D.C., Samocha, K.E., Barclay, K., Kolk, M., Niemi, M.E.K., Kirov, G., Martin, H.C., and Hurles, M.E. (2020). Sex-biased reduction in reproductive success drives selective constraint on human genes (Genetics).

23. Goodrich, J., Singer-Berk, M., Son, R., Sveden, A., Wood, J., England, E., Cole, J.B., Weisburd, B., Watts, N., Zappala, Z., et al. Determinants of penetrance and variable expressivity in monogenic metabolic conditions across 77,184 exomes. 31.

24. Tuke, M.A., Ruth, K.S., Wood, A.R., Beaumont, R.N., Tyrrell, J., Jones, S.E., Yaghootkar, H., Turner, C.L.S., Donohoe, M.E., Brooke, A.M., et al. (2019). Mosaic Turner syndrome shows reduced penetrance in an adult population study. Genet. Med. 21, 877–886.

25. Landrum, M.J., Lee, J.M., Benson, M., Brown, G., Chao, C., Chitipiralla, S., Gu, B., Hart, J., Hoffman, D., Hoover, J., et al. (2016). ClinVar: public archive of interpretations of clinically relevant variants. Nucleic Acids Res. 44, D862–D868.

26. Fry, A., Littlejohns, T.J., Sudlow, C., Doherty, N., Adamska, L., Sprosen, T., Collins, R., and Allen, N.E. (2017). Comparison of Sociodemographic and Health-Related Characteristics of UK Biobank Participants With Those of the General Population. Am. J. Epidemiol. 186, 1026–1034.

27. Plomin, R., and Deary, I.J. (2015). Genetics and intelligence differences: five special findings. Mol. Psychiatry 20, 98–108.

